# Clinical guideline variability in the diagnosis and care of people at risk for hereditary hematopoietic malignancy syndromes

**DOI:** 10.1101/2023.04.20.23288424

**Authors:** Adam Hamidi, Gregory W. Roloff, Reid Shaw, Maria Acevedo, Shaili Smith, Michael W. Drazer

**Author notes:** Correspondence: Michael W. Drazer, MD, PhD, Section of Hematology/Oncology, The University of Chicago, 5841 S. Maryland Ave, MC 2115, Chicago, IL 60637. Contributed equally (Adam Hamidi, Gregory W. Roloff).

## Abstract

A growing understanding of the complexities of hematopoietic malignancies necessitates the existence of clinical recommendations that are sufficiently comprehensive. Although hereditary hematopoietic malignancies (HHMs) are increasingly recognized for conferring risk of myeloid malignancy, frequently utilized clinical recommendations have never been appraised for the ability to reliably guide HHM evaluation. We assessed established society-level clinical guidelines for inclusion of critical HHM genes and graded the strength of testing recommendations. We uncovered a substantial lack of consistency of recommendations guiding HHM evaluation. Such heterogeneity in guidelines likely contributes to refusal by payers to support HHM testing, leading to underdiagnoses and lost opportunities for clinical surveillance.

---

A substantial proportion of blood cancers are now known to be hereditary hematopoietic malignancies (HHMs) driven by germline variants that adhere to Mendelian inheritance patterns. Approximately 14% of older patients with acute myeloid leukemia (AML), for example, carry HHM-associated germline variants [1]. Similarly, at least 7% of patients undergoing stem cell transplant for myelodysplastic syndrome (MDS) carry germline variants associated with hereditary hematopoietic disorders [2]. The prevalence of HHMs (7-14%) in relatively unselected groups of patients is similar to the prevalence of other biologically relevant subgroups of MDS/AML, such as *FLT3*-mutated AML (approximately 30%), *IDH1*-mutated MDS/AML (10%), and *IDH2*-mutated MDS/AML (10%) [3]. The clinical recognition of patients with HHMs is crucial so as to avoid donor-derived cancer, to counsel family members regarding genetic testing, and to identify potential treatment options [2,4]. Given the common prevalence of HHMs, there is an urgent need to develop consistent standards for the diagnosis and care of patients at risk for HHMs. For example, we previously showed next generation sequencing-based HHM assays are technically inadequate to diagnose many HHMs [5,6]. In our experience, diagnostic germline testing for HHMs is also frequently denied by third-party payers. Frequent insurance denials have also occurred in other hereditary cancer syndromes [7].

Guidelines from organizations such as the National Comprehensive Cancer Network (NCCN), the American Society of Clinical Oncology (ASCO), and similar groups are often used to support decisions regarding third party reimbursement of diagnostic assays. The degree to which current clinical guidelines support the evaluation of patients with possible HHMs, however, has not been evaluated. Heterogeneity in these guidelines could inadvertently lead payers to deny coverage for medically indicated HHM evaluations. To address this knowledge gap, we analyzed clinical guidelines from all groups with published recommendations for MDS and/or AML. We then determined the heterogeneity in HHM-related recommendations from these groups.

Eight sets of clinical guidelines for MDS or AML were analyzed. We excluded publications focused solely on pathologic classifications, such as those from the World Health Organization, as these are rarely used to justify third party payment decisions. We determined if each guideline included criteria regarding the diagnosis and evaluation of patients at risk for HHMs. For guidelines that discussed HHMs, we then determined which genes, if any, were recommended for germline sequencing. The strength of each recommendation was determined using a scale: “Not Addressed” (no mention of HHMs), “Consider Testing” (HHMs discussed, but without clear testing criteria), or “Firm Recommendation” (clear criteria for HHM testing provided). We used R version 4.2.2 to generate heat maps of genes in each publication and the strength of recommendation from each group.

The most up-to-date clinical guidelines for MDS were from the NCCN Clinical Practice Guidelines in Oncology: Myelodysplastic Syndromes v.1.2023 [8], the European Society of Medical Oncology (ESMO) Clinical Practice Guidelines for diagnosis, treatment and follow-up [9], and the British Society of Haematology guidelines for the diagnosis and evaluation of prognosis of adult myelodysplastic syndromes [10]. NCCN guidelines discussed the largest number of HHM-related genes (n=45) and made clear clinical recommendations in terms of eligibility criteria for HHM testing. ESMO suggested that clinicians “consider testing” eight HHM-related genes. The BSH suggested that clinicians consider testing three genes (**Figure 1**).

**Figure 1.**
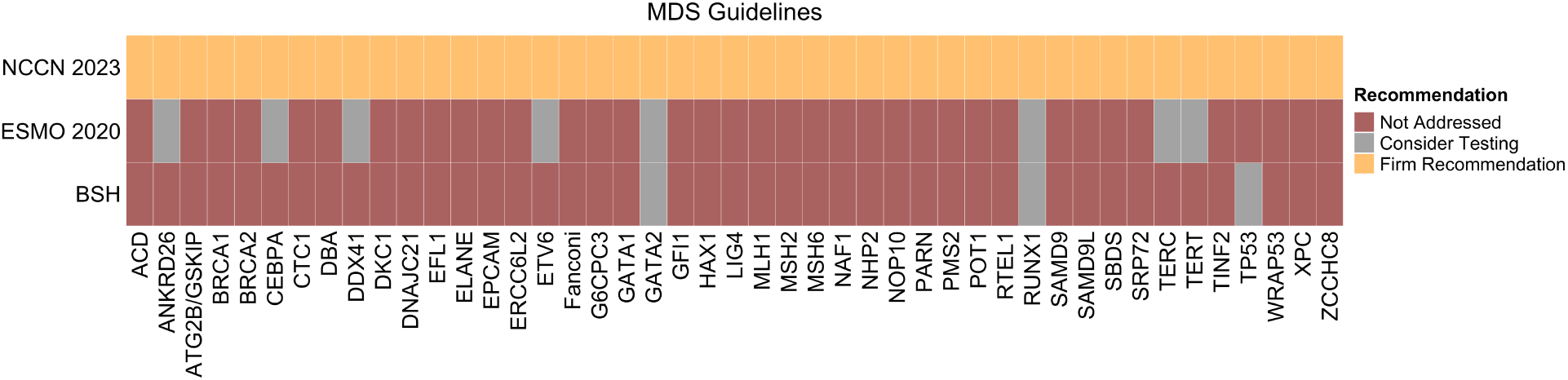
Recommendations for HHM-focused evaluation of patients with MDS across clinical guidelines. Genes included for HHM evaluation are on the horizontal axis. Recommendations were scaled based on the strength of the language used. “Fanconi” refers to the full spectrum of Fanconi anemia genes. “DBA” refers to the full spectrum of Diamond Blackfan anemia genes.

Five clinical guidelines for AML were identified: the NCCN Clinical Practice Guidelines in Oncology: Acute Myeloid Leukemia v.3.2022 [11], the ESMO 2020 Clinical Practice Guidelines for diagnosis, treatment and follow-up [12], the ASCO initial diagnostic workup of acute leukemia [13], the European LeukemiaNet (ELN) 2022 update [14], and the Nordic recommendations for the genetic diagnosis, clinical management, and follow-up of germline predisposition to myeloid neoplasms [15]. A total of 64 genes were included across these guidelines. The Nordic guidelines included the largest number of genes (41) with clear testing criteria. The NCCN provided clear criteria for thirteen genes. Both the ELN and ESMO guidelines suggested testing for select HHM-related genes without clear criteria. While mentioning the importance of HHMs, ASCO did not discuss testing criteria and did not include specific HHM-related genes (**Figure 2**).

**Figure 2.**
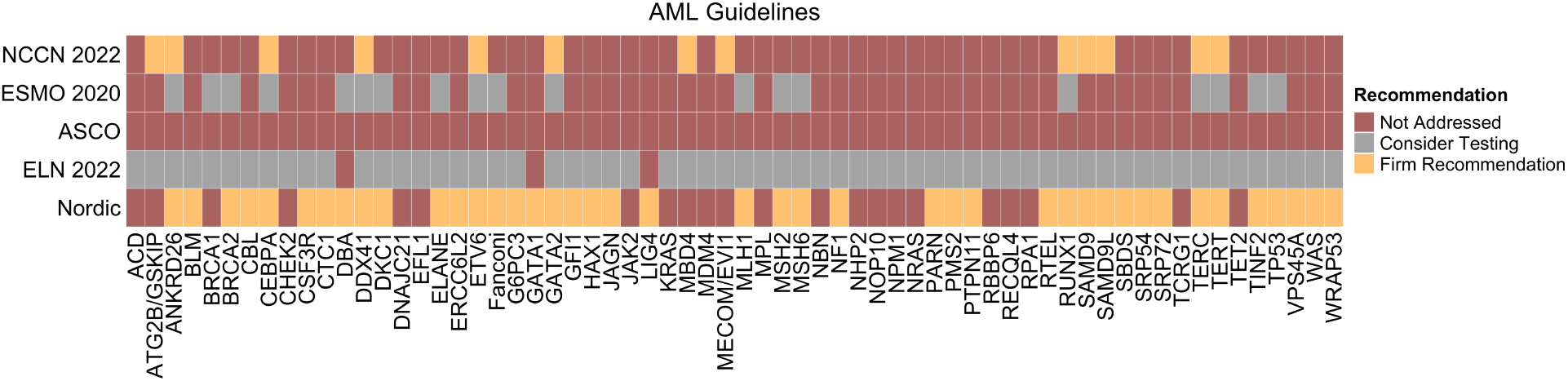
Recommendations for HHM-focused evaluation of patients with AML across clinical guidelines. Genes included for HHM evaluation are on the horizontal axis. Recommendations were scaled based on the strength of the language used. “Fanconi” refers to the full spectrum of Fanconi anemia genes. “DBA” refers to the full spectrum of Diamond Blackfan anemia genes.

Rapid advances in the biological understanding of blood cancers and in the clinical care of people with these diseases have necessitated the development of updated clinical guidelines. HHMs, many of which have been discovered only in the past decade, are a “case study” in the rapid pace of the scientific understanding of the genetic origins of MDS and AML. These advances necessitate the development of up-to-date guidelines that reflect the most contemporary developments in the clinical care of people with MDS and AML.

Here, we performed the first analysis of HHM-specific recommendations in clinical guidelines of the care of people with MDS and/or AML. Our analysis revealed marked heterogeneity and inconsistency in recommendations regarding HHM diagnosis and testing. This heterogeneity may potentially lead to denials of coverage by third party payers, which could frustrate clinicians and make them reticent to pursue HHM evaluations. Inconsistent clinical guidelines, therefore, may ultimately lead to HHM underdiagnosis and less optimized care of patients with these syndromes. Harmonizing and updating clinical MDS/AML guidelines to include the full spectrum of HHM-related variants, as well as the inclusion of clear eligibility criteria for HHM testing, will facilitate the accurate diagnosis and care of patients with these syndromes.

## Data Availability

All data produced in the present study are available upon reasonable request to the authors

## Notes

### Competing Interest Statement

The authors have declared no competing interest.

### Funding Statement

This study did not receive any funding

## Bibliography & References Cited

1. Yang F, Long N, Anekpuritanang T, et al. Identification and prioritization of myeloid malignancy germline variants in a large cohort of adult patients with AML. Blood. 2022 Feb 24;139(8):1208–1221.

2. Feurstein S, Trottier AM, Estrada-Merly N, et al. Germ line predisposition variants occur in myelodysplastic syndrome patients of all ages. Blood. 2022 Dec 15;140(24):2533–2548.

3. Ahmadmehrabi K, Haque AR, Aleem A, et al. Targeted Therapies for the Evolving Molecular Landscape of Acute Myeloid Leukemia. Cancers (Basel). 2021 Sep 16;13(18).

4. Roloff GW, Drazer MW, Godley LA. Inherited Susceptibility to Hematopoietic Malignancies in the Era of Precision Oncology. JCO Precis Oncol. 2021 Nov;5:107–122.

5. Roloff GW, Godley LA, Drazer MW. Assessment of technical heterogeneity among diagnostic tests to detect germline risk variants for hematopoietic malignancies. Genet Med. 2021 Jan;23(1):211–214.

6. Roloff GW, Shaw R, O’Connor TE, et al. Stagnation in quality of next-generation sequencing assays for the diagnosis of hereditary hematopoietic malignancies. J Genet Couns. 2023 Jan 15.

7. Amendola LM, Hart MR, Bennett RL, et al. Insurance coverage does not predict outcomes of genetic testing: The search for meaning in payer decisions for germline cancer tests. J Genet Couns. 2019 Dec;28(6):1208–1213.

8. National Comprehensive Cancer Network. Myelodysplastic Syndromes, Version 1.2023, NCCN Clinical Practice Guidelines in Oncology. 2022.

9. Fenaux P, Haase D, Santini V, et al. Myelodysplastic syndromes: ESMO Clinical Practice Guidelines for diagnosis, treatment and follow-up(dagger☆). Ann Oncol. 2021 Feb;32(2):142–156.

10. Killick SB, Wiseman DH, Quek L, et al. British Society for Haematology guidelines for the diagnosis and evaluation of prognosis of Adult Myelodysplastic Syndromes. Br J Haematol. 2021 Jul;194(2):282–293.

11. National Comprehensive Cancer Network. Acute Myeloid Leukemia, Version 3.2022, NCCN Clinical Practice Guidelines in Oncology. 2023.

12. Heuser M, Ofran Y, Boissel N, et al. Acute myeloid leukaemia in adult patients: ESMO Clinical Practice Guidelines for diagnosis, treatment and follow-up. Ann Oncol. 2020 Jun;31(6):697–712.

13. de Haas V, Ismaila N, Advani A, et al. Initial Diagnostic Work-Up of Acute Leukemia: ASCO Clinical Practice Guideline Endorsement of the College of American Pathologists and American Society of Hematology Guideline. J Clin Oncol. 2019 Jan 20;37(3):239–253.

14. Dohner H, Wei AH, Appelbaum FR, et al. Diagnosis and management of AML in adults: 2022 recommendations from an international expert panel on behalf of the ELN. Blood. 2022 Sep 22;140(12):1345–1377.

15. Baliakas P, Tesi B, Wartiovaara-Kautto U, et al. Nordic Guidelines for Germline Predisposition to Myeloid Neoplasms in Adults: Recommendations for Genetic Diagnosis, Clinical Management and Follow-up. Hemasphere. 2019 Dec;3(6):e321.

